# Development and validation of a LC-MS/MS method for ripretinib and its metabolite: example of a journey from laboratory bench to routine application with a greenness assessment

**DOI:** 10.1101/2025.04.03.25325083

**Authors:** Cedric Rakotovao, Ahmad Sharanek, Audrey Burban, Paul Gueroue, Stéphane Bouchet, Guillaume Bouguéon, Dominique Ducint, Mathieu Molimard, Antoine Italiano, Sarah Djabarouti, Joris Guyon

## Abstract

Therapeutic drug monitoring of protein kinase inhibitors is a widely practiced worldwide. Based on the example of ripretinib dosage requested by a clinician, we detailed the process of method development, using a literature-based approach while ensuring the sustainability of the method to be as environmentally friendly as possible. Therefore, an UPLC-MS/MS method for ripretinib and its active metabolite was optimized and validated using the corresponding stable isotopic internal standards in human plasma. The procedure has employed mobile phase mixture of water with 1% acetic acid and 0.1% formic acid, and acetonitrile. Positive electrospray ionization was performed coupling with multiple reaction monitoring of m/z 510.4→417.4 and 510.4→389.4 for ripretinib, and 496.3→403.3 and 496.3→375.3 for N-desmethyl-ripretinib. The method was successfully validated according the current version of ICH Guideline provided by the EMA. The greenness assessment score of this procedure was evaluated and compared with previously published approaches using the AGREE metric. The validated UPLC-MS/MS method successfully monitored ripretinib and its metabolite concentrations in clinical and pre-clinical models.

## 1 Introduction

Therapeutic drug monitoring (TDM) is an essential biological service in hospitals, particularly for drugs with a narrow therapeutic index, pharmacokinetic variability or significant toxicity (1).

Plasma drug concentrations are measured using various analytical techniques, selected based on the properties of the drug, concentration levels, cost, available equipment, and sample type. Two main analytical method can be distinguished: immunochemical and chromatographic methods. The first method uses an antibody specific to the molecule to be assayed and can be rapid, especially if a commercial kit is available for automation. If this is not the case, laboratories generally use the second method, which analyzes a mixture by separating its individual components. This separation can be carried out using gas or liquid chromatography (LC), depending on the characteristics of the molecules to be assayed. It requires a preliminary sample preparation step, such as extraction or precipitation, and is coupled with detection methods like UV detection, mass spectrometry (MS), or flame ionization detection. In the latter case, it is necessary for each laboratory to validate its assay method in order to determine its performance and limits (2).

Laboratories are also encouraged to adopt sustainable development measures. Green analytical chemistry has therefore received particular attention in recent years, focusing on the reduction of harmful chemicals, energy consumption and waste in analytical procedures (3,4). Numerous metric approaches are available to assess the greenness of a method (5). Analytical Greenness Metric Approach (AGREE) was mainly used to assess greening, both because of its ease of use and because it is based on 12 principles of green analytical chemistry (6).

In this study, we present the various steps taken by the laboratory in response to a request of a clinician for ripretinib plasma monitoring. Ripretinib (or DCC-2618) is the latest drug to be approved for the treatment of gastrointestinal stromal tumor (GIST), a rare type of cancer that originates in the connective tissues of the gastrointestinal tract (7,8). Ripretinib and its main active metabolite, N- desmethyl-ripretinib (DP-5439), inhibit tumor growth of primary and drug-resistant KIT and PDGFRA mutants (9).

These molecules have a safety profile similar to that of other protein kinase inhibitors (PKI), including serious side effects such as hypertension or cardiac dysfunction (10–12). Moreover, ripretinib and N-desmethyl-ripretinib also inhibit the activity of VEGFR2 and could favour conditions conductive to arterial dissections or aneurysms (13,14).

Over 99% of ripretinib and N-desmethyl-ripretinib are bound to plasma proteins, their metabolism is mainly dependent on cytochrome p450 3A4/5, and their pharmacokinetics appear to show a non-linear relationship with doses above 150 mg (8). These parameters lead to highly variable drug exposure from one patient to another, resulting in plasma concentrations that are often too high or too low. In addition, ripretinib pharmacokinetics were altered by the administration of CYP450 inhibitors or inducers (15,16). Therefore, TDM of these molecules may be advisable to assist clinicians.

Here, we proposed a LC-MS/MS method for ripretinib and its metabolite based on a scientific literature analysis, emphasizing the evaluation of green analytical chemistry. This is followed by technical and biological validation in accordance with the current version of the ICH guidelines on bioanalytical method validation provided by the EMA (17). Finally, the method will be tested on both clinical and preclinical models.

## 2 Results and discussion

### 2.1 Literature review and greenness analysis

All information about ripretinib and/or N-desmethyl-ripretinib analysis is recorded in **Table 1** (15,16,18,19). The previous analyses utilized either the precipitation technique for sample preparation or liquid-liquid extraction. The pre-clinical study by Wang *et al*. is optimized for greater sensitivity (LLOQ at 1 ng/mL) but involves a number of steps, including evaporation to concentrate the sample. The method by Lin *et al*. reported the widest measurement range, with concentrations ranging from 5 to 5000 ng/mL. They also offered the advantage of requiring significantly fewer preparation steps. However, its analysis time is considerably longer—6 minutes compared to 2 minutes for the two methods that quantify only ripretinib. All studies use one quantitative product ion for ripretinib quantification, approximately 417 m/z for Wang, Qian and Lin studies., and 94 m/z for Mudawath study.

**Table 1.** Comparison of analytical LC-MS/MS method parameters found in the literature.

|  | Wang et al., 2021 | Mudavath et al., 2023 | Qian et al., 2024 | Lin et al., 2025 | Developed method |
| --- | --- | --- | --- | --- | --- |
| **Instrumentation** |  |  |  |  |  |
| Stationary phase | ACQUITY UPLC BEH C18 column (2.1 mm × 50 mm, 1.7 μm) | Zorbax SB C18 column (4.6 mm × 50 mm, 3.5 μm) | ACQUITY UPLC HSS T3 column (2.1 mm × 50 mm, 1.8 μm) | ThermoFisher Hypersil GOLDTM C18 HPLC column (4.6 mm × 50 mm, 5 μm) | Acquity Cortecs C18 column (2.1 × 50 mm, 1.6 μm) |
| Mobile phases | A: 0.1% formic acid<br>B: acetonitrile | A: formic acid and 10 mM ammonium formate<br>B: acetonitrile | A: 0.1% formic acid and 5 mM ammonium formate<br>B: acetonitrile | A: 0.1% formic acid<br>B: acetonitrile | A: 1% acetic acid and 0.1% formic acid<br>B: acetonitrile |
| Flow rate | 0.4 mL/min | 0.4 mL/min | 0.5 mL/min | 0.5 mL/min | 0.3 mL/min |
| Mass spectrometer | Waters Xevo TQ-S | Sciex API-3000 | Sciex QTRAP 4500 | Agilent 6420 | Waters Acquity TQD |
| Ionization source | ESI | ESI | ESI | ESI | ESI |
| Analyzed molecule | **Ripretinib** | **Ripretinib** | **Ripretinib**<br>**N-d-ripertinib** | **Ripretinib**<br>**N-d-ripertinib** | **Ripretinib**<br>**N-d-ripertinib** |
| Transitions (m/z) | 509.93→416.85 | 510.09→94.06 | **Ripretinib:** 510.1→417.1<br>**N-d-ripertinib:** 496.0→403.1 | **Ripretinib:** 510.1→417<br>**N-d-ripertinib:** 496.11→402.9 | **Ripretinib:** 510.4→417.4 and 510.4→389.4<br>**N-d-ripertinib:** 496.3→403.3 and 496.3→375.3 |
| Time of analysis | 2 min | 2 min | 4.7 min | 6 min | 3 min |
| **Method** |  |  |  |  |  |
| Matrix | Beagle Dog | Wistar Rat | Human | Human | Human and mouse |
| Internal standard | avapritinib | imatinib | [<sup>2</sup>H<sub>8</sub>]-ripertinib | [<sup>2</sup>H<sub>8</sub>]-ripertinib | [<sup>13</sup>C<sub>6</sub>]-ripertinib<br>[<sup>13</sup>C<sub>6</sub>]-N-d-ripertinib |
| Sample preparation | From 100 μL of matrix<br>- addition 20 μL of IS<br>- addition 200 μL sodium hydroxide<br>- addition 1 mL ethyl acetate<br>- vortex at 24°C and evaporation with nitrogen<br>- addition 100 μL mobile phase and transfer into a vial | From 500 μL of matrix<br>- addition 50 μL of IS<br>- addition 100 μL 0.1% formic acid<br>- Liquid-liquid extraction with 2 × 1.5 mL dichloromethane<br>- organic layer flash-frozen<br>- evaporation with nitrogen<br>- addition 500 μL mobile phase and transfer into a vial | From 50 μL of matrix<br>- addition 200 μL of ACN containing IS<br>- vortex and centrifugation at 4°C<br>- transfer 10 μL of supernatant into a vial with 90 μL CAN<br>- vortex and mixed | From 100 μL of matrix<br>- addition 5 μL of IS<br>- addition 300 μL of ACN<br>- vortex and centrifugation at 4°C<br>- transfer of supernatant into a vial | From 50 μL of matrix<br>- addition 150 μL of ACN containing IS using a repeating pipette<br>- vortex and centrifugation at 24°C<br>- transfer of supernatant into a vial |
| Linearity (ng/mL) | 1 to 1000 | 20 to 1000 | **Ripretinib:** 7.5 to 3000<br>**N-d-ripertinib:** 10 to 4000 | **Ripretinib:** 10 to 5000<br>**N-d-ripertinib:** 10 to 5000 | **Ripretinib:** 50 to 5000<br>**N-d-ripertinib:** 50 to 5000 |

Abbreviation: ACN, Acetonitrile; ESI, Electron spray ionization; MS, Mass Spectrometry

Wang and Mudavath studies used avapritinib and imatinib as internal standard (IS), respectively. The two most recent methods used [^2^H_8_]-ripretinib as IS. The use of an isotopic internal standard is essential for accurate quantification, as it often compensates for matrix effects or variations in recovery. However, reproducibility is not always optimal when using [^2^H]-IS and when used, it is better suited to have an IS with fewer [^2^H] (20–22).

For the comparison of analytical procedure greenness, the four methods were evaluated using the AGREE tool (6). The overall results are shown in **Figure 1**. The sample pretreatment is external and involves a reduced number of steps (Principle 1). A volume of 0.05 mL plasma is needed for Qian study, 0.1 mL plasma is required for Wang study and Lin study while 0.5 mL plasma is needed for Mudavath study (P2). The measurements are off-line (P3). The procedure involves many distinct steps for Mudavath study and fewer for Wang and Lin studies (P4). Procedures are manual and involve a miniaturized sample preparation technique (P5). No derivatization agents are involved (P6). The total waste generated, including extraction or precipitation solvents, laboratory plastics (tubes, vials, etc.), and the mobile phases, is approximately 6.1 g for Wang study, 8.8 g for Mudavath study, 5.1 g for Qian study, and 5.8 g for Lin study (P7). Only ripretinib are determined in a single run for Wang and Mudavath studies, while both ripretinib and its metabolite are determined for Qian and Lin studies. The sample throughput is 30 samples per hour for Wang and Mudavath studies, 12.8 samples per hour for Qian study and 10 samples per hour for Lin study (P8). LC-MS is the most energy-consuming technique (P9). None of the reagents are derived from bio-based sources (P10). Procedures require approximately 2.2 mL, 4.4 mL, 2.55 mL, and 3.3 mL of toxic solvents for Wang, Mudavath, Qian and Lin studies, respectively (P11). Threats not avoided include highly flammable and corrosive substances (P12). The scores for the procedures obtained with AGREE assessment are 0.43, 0.51, 0.52 and 0.54 for Mudavath, Lin, Wang et Qian studies, respectively.

**Figure 1:**
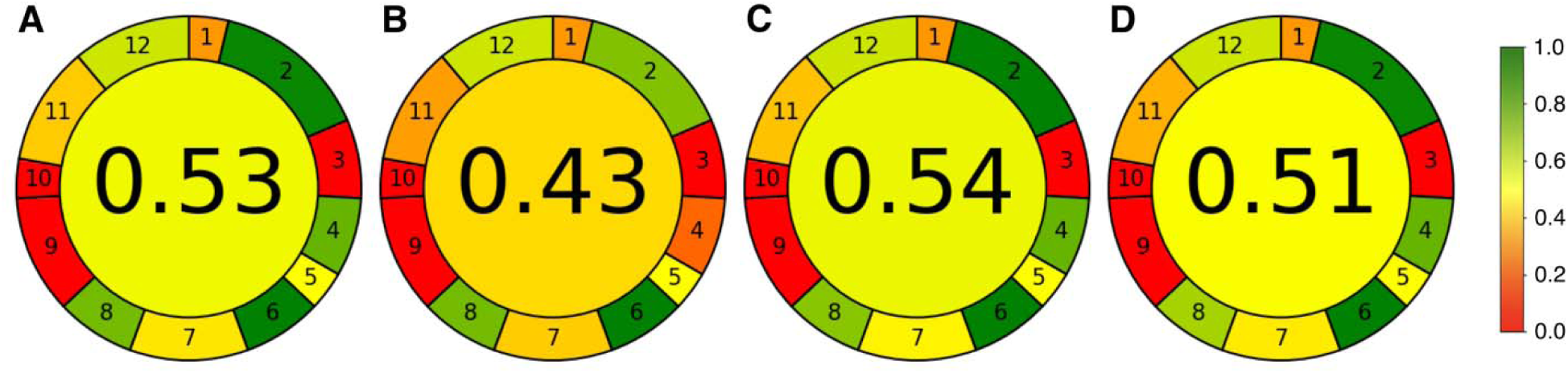
Results of AGREE analysis for literature review procedures. Generic greenness assessment results for (**A**) Wang study, (**B**) Mudavath study, (**C**) Qian study and (**D**) Lin study. The overall score is displayed in the middle of each circle.

### 2.2 Method development

The expertise of a laboratory and its analytical equipment will guide the choice of the analytical method to be used. For TKI quantification, as presented in the literature, we have chosen LC-MS/MS, a sensitive and specific method that is particularly well-suited for the quantification of small molecules. Moreover, the respective [^13^C_6_]-IS were used, as they were best suited for LC-MS/MS quantification (20,21).

The first step involves injecting a pure solution of the compound into the LC-MS/MS and scanning an m/z range that includes the expected m/z value of the compound. Chemical structures of ripretinib, N-desmethyl-ripretinib are shown in **Figure 2**. Both compounds contain a bromine atom with a molecular structure composed of two naturally occurring isotopes, ^79^Br and ^81^Br, which are present in approximately equal natural abundance of 50% each. This explains the unique mass spectrum of the two compounds, characterized by two peaks of similar intensity: 510.4 m/z and 512.4 m/z for ripretinib and 496.3 m/z and 498.3 m/z for N-desmethyl-ripretinib.

**Figure 2:**
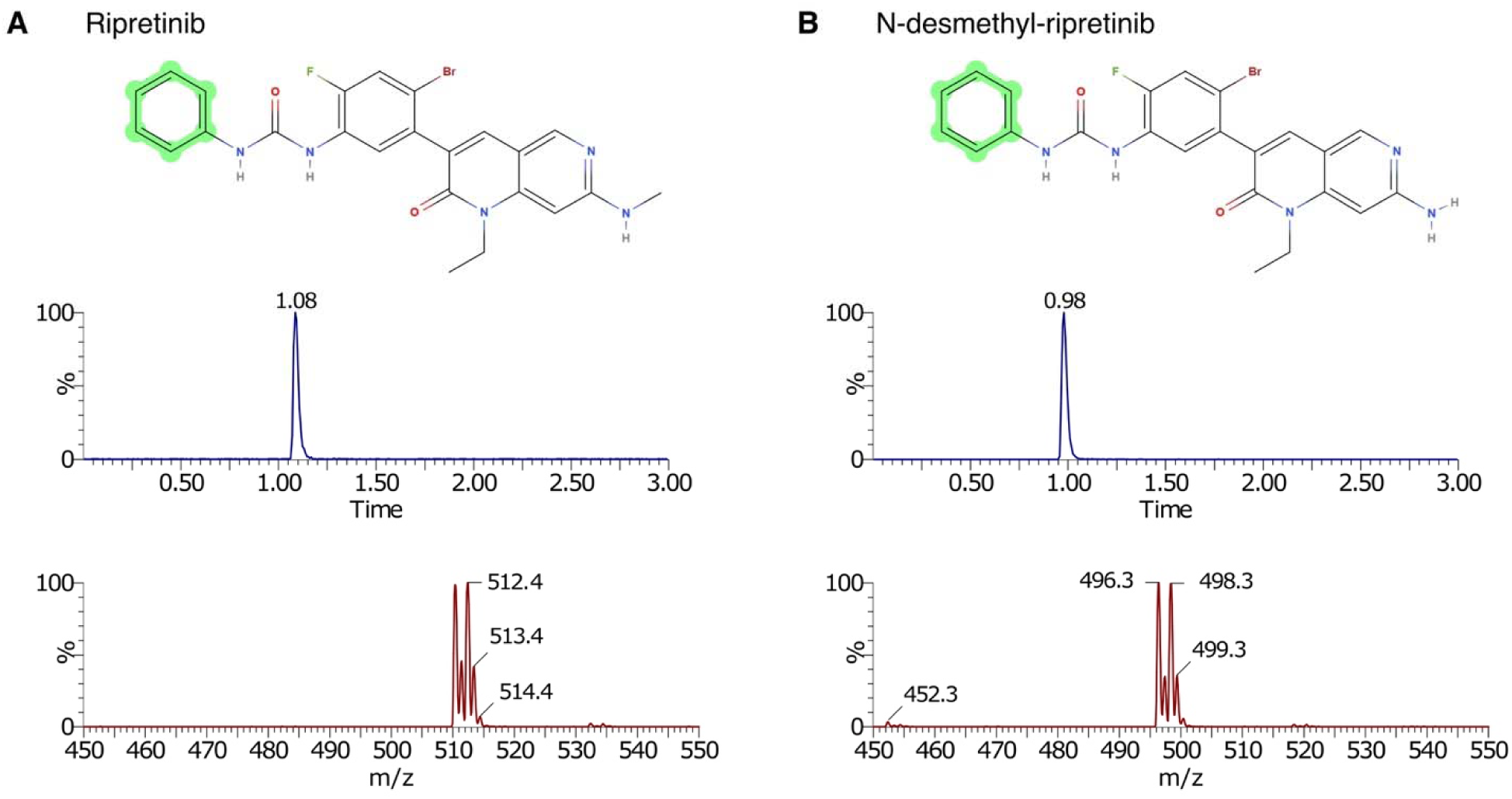
Chromatograms and Mass Spectra of Ripretinib and N-desmethyl-ripretinib. Chemical structures, representative chromatograms (*blue*) and mass spectra (*red*) of (**A**) ripretinib and (**B**) N-desmethyl-ripretinib. Benzene highlighted in green represents carbons that have been replaced by ^13^C to form the corresponding stable isotope. Chemical structure was drawn using Molview (https://molview.org/).

Once the m/z is determined, we optimized the signal (abundance) of the compound by adjusting the cone voltage. It is important to note that a high voltage can degrade the compounds, leading to a decreased signal. As shown in **Figure 3A** and **B**, the optimal voltage under our analytical conditions was 50V. LC-MS/MS speeds up selected reaction monitoring by scanning for both precursor ions (the compound of interest) and product ions. This approach enhances sensitivity by allowing longer monitoring of specific fragment ions. The first mass analyzer selects a specific precursor ion, while the second scans for all its fragmented ions (“products”) in the collision cell. The quantification product-ions were 510.4→417.4 and 496.3→403.3, while the confirmation product-ion were 510.4→389.4 and 496.3→375.3 for ripretinib and N-desmethyl-ripretinib, respectively (**Figure 3C** and **D**). We introduced confirmation product-ions to enhance the specificity of the method compared to previous studies (**Table 1**). All mass spectrometer parameters for each analyte, including respective IS, are displayed in **Table 2**.

**Figure 3.**
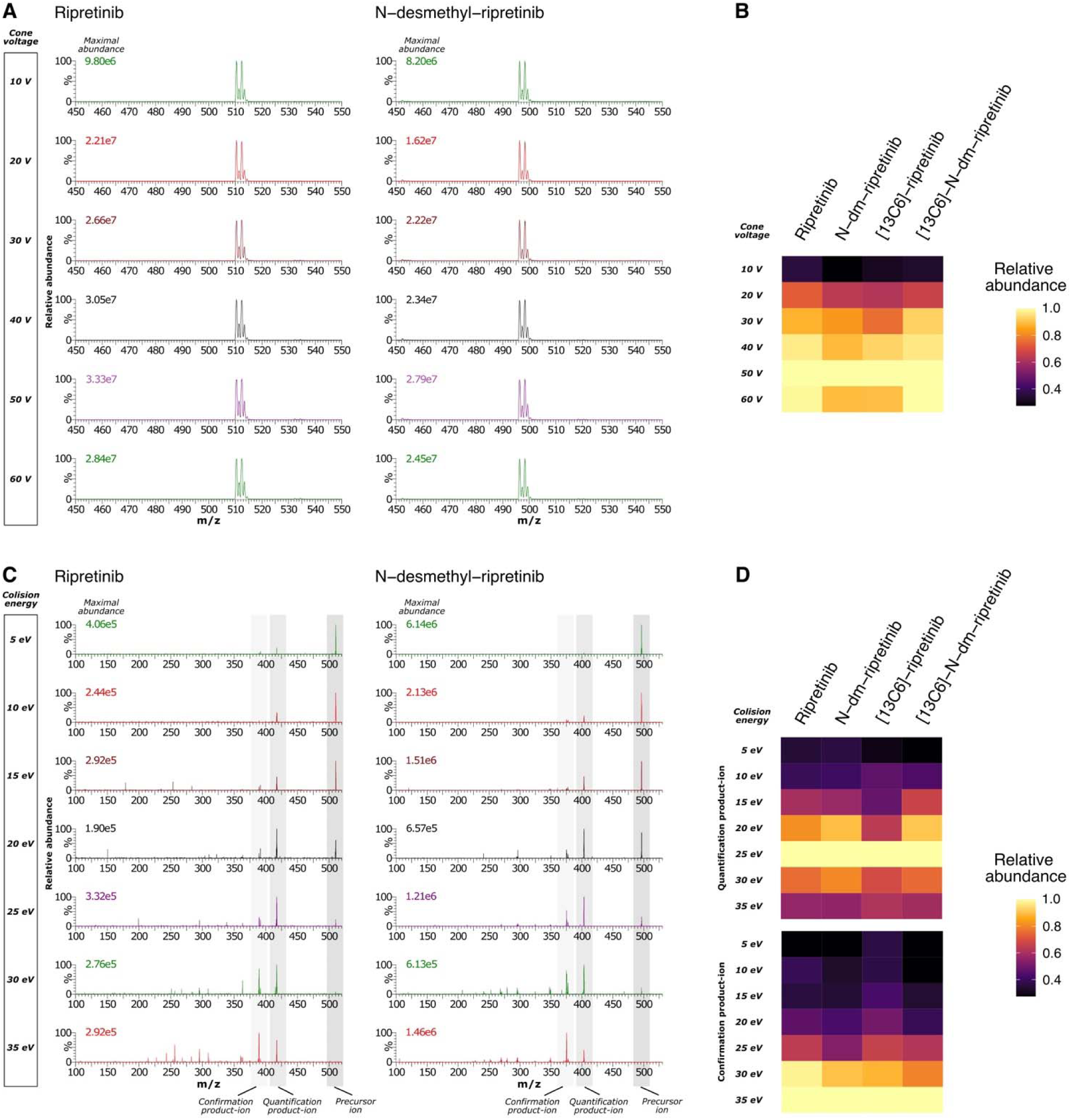
Mass spectrometer settings determination. (**A**) and (**B**) Cone voltage determination. (**A**) Mass spectra of ripretinib and N-desmethyl-ripretinib showing the maximal abundance of the compounds according the different cone voltage (one representing injection). (**B**) Heatmap showing the distribution of ripretinib, N-desmethyl-ripretinib and the respective IS (3 independent injections). The observed abundance is relative to the maximal abundance observed for each compound. (**C**) and (**D**) Product-ion determination. (**C**) Mass spectra of ripretinib and N-desmethyl-ripretinib showing the fragmentation of compounds according the different collision energy (one representing injection). (**D**) Heatmap showing the distribution of the quantification product-ion and confirmation product-ion of ripretinib, N-desmethyl-ripretinib and the respective IS (3 independent injections). The observed abundance is relative to the maximal abundance observed for each compound.

**Table 2.** Mass spectrometer settings for the detection of ripretinib and N-desmethyl-ripretinib by UPLC-MS/MS.

| Drugs | Retention time (min) | Precursor ion (m/z) | Cone voltage (V) | Quantification product-ion (m/z) | Collision energy (eV) | Confirmation product-ion (m/z) | Collision energy (eV) |
| --- | --- | --- | --- | --- | --- | --- | --- |
| ripretinib | 1.08 | 510.4 | 50 | 417.4 | 25 | 389.4 | 35 |
| [13C6]-ripretinib | 1.08 | 516.4 | 50 | 417.4 | 25 | 389.4 | 35 |
| DP-5439 | 0.98 | 496.3 | 50 | 403.3 | 25 | 375.3 | 35 |
| [13C6]-N-desmethyl-ripretinib | 0.98 | 502.3 | 50 | 403.3 | 25 | 375.3 | 35 |

The criteria for lower environmental impact, on which a laboratory can play a more flexible role, correspond to optimization of the waste generated, depending on the intake volume, the type of extraction, number of steps, analysis time and chromatographic throughput. Of course, the use of green solvents could also be considered (23), which was not the case here. Based on the previous literature review, the effects of different stationary phases, mobile phase compositions, gradient elution and flow rates were studied in order to optimize chromatographic conditions in a more environmentally friendly way (**Table 1**). The analytical run time was 3 min, and the retention time was 1.08 min for ripretinib and 0.98 min for N-desmethyl-ripretinib (**Figure 1**).

### 2.3 Method validation

Method validation is then conducted in accordance with the current ICH guidelines on bioanalytical method validation provided by the EMA (**Supplementary Table S1**) (17).

#### 2.3.1 Matrix Effect

The matrix effect is tested on several types of plasma, including human and mouse plasma. No signal alteration was observed for ripretinib and N-desmethyl-ripretinib. In conclusion, the current research does not provide evidence of a significant influence of various matrices.

#### 2.3.2 Selectivity and Specificity

Injection of six individual drug-free human plasmas (including one with hemolysis and one with hyperlipidemia) into the MRM channels of ripretinib and N-desmethyl-ripretinib revealed no interference from endogenous substances (**Supplementary Figure S1A-B**). Injection of six individual human plasmas spiked with various compounds revealed no interference from exogenous molecules, including various PKI (**Supplementary Figure S1C-D**). The abundance of ripretinib and N-desmethyl-ripretinib was reported to be less than 20% of the LLOQ, and their respective IS abundances were below 5% (**Supplementary Figure S1B-D**). The results showed that the method was selective and specific for ripretinib and N-desmethyl-ripretinib, as all compounds satisfied the predetermined acceptance criteria.

#### 2.3.3 Linearity

The LLOQ for ripretinib and its metabolite N-desmethyl-ripretinib was determined to be 50 ng/mL according to the plasma concentration in clinical trials (11,24) and **Supplementary Figure S2**. The calibration curves followed a linear regression with a 1/x² weighted least-squares model (**Figure 4**). R² was higher than 0.999 for ripretinib and N-desmethyl-ripretinib within the concentration ranging from 50 to 5000 ng/mL (LLOQ and ULOQ). The resulting average equation for ripretinib was *y* = 6.67 × 10^−4^*x* + 0.0024 and for N-desmethyl-ripretinib *y* = 6.90 × 10^−4^*x* + 0.0047. The concentrations of the calibration standards differed from the experimental concentration by ±11%. The calibration met the linearity requirement, in line with established criteria.

**Figure 4.**
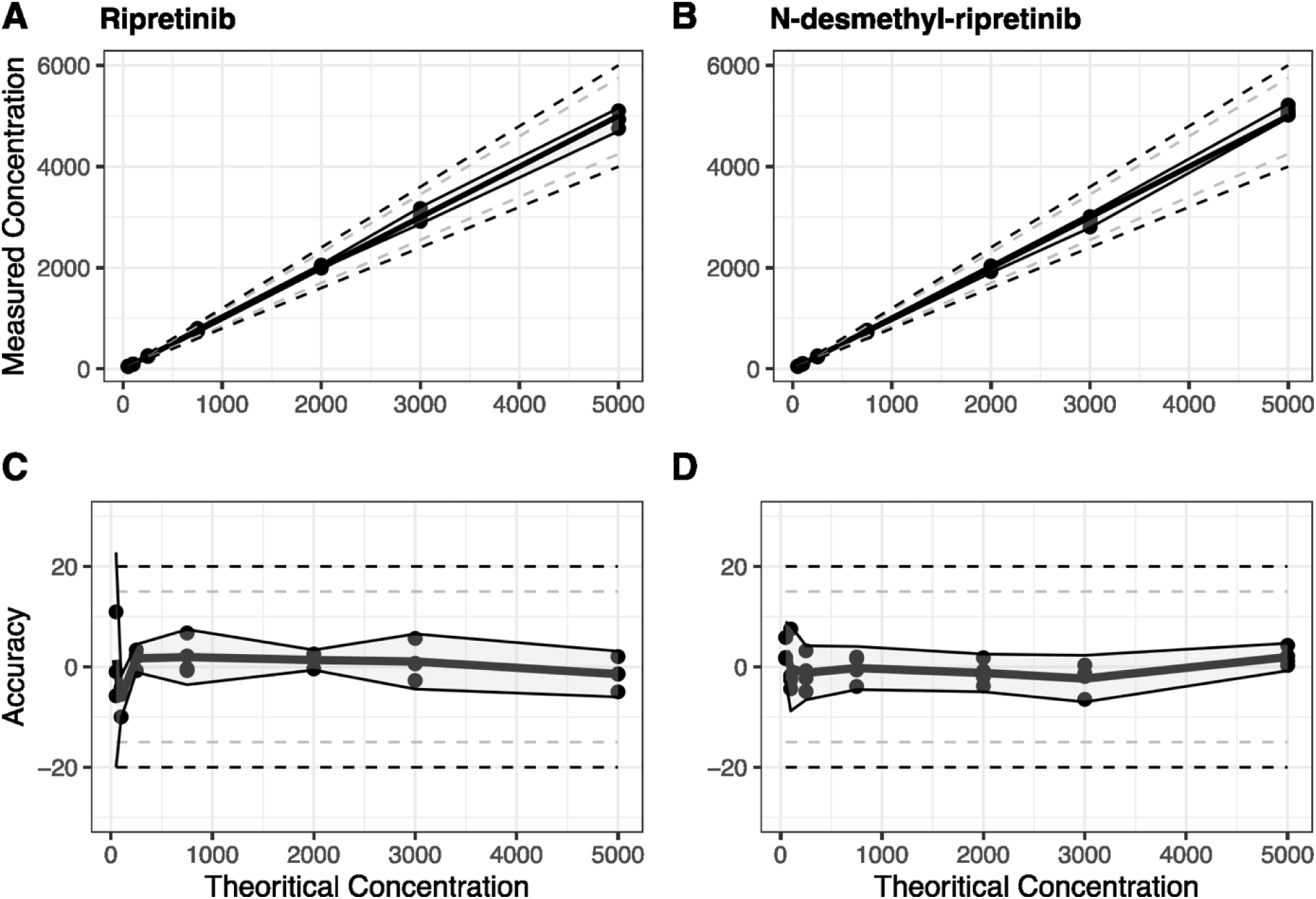
Linearity analysis. (**A**) Calibration curve and (**B**) residual plot resulting from linear regression with a 1/x² weighted least-square model of ripretinib (*left*) and N-desmethyl-ripretinib (*right*) (n = 4). Grey dashed lines represent a variation of ± 15% and black dashed lines a variation of ± 20%. The interval around the values corresponds to the 95% confidence interval with k=1.96.

#### 2.3.4 Repetitive and intermediate fidelity

Intra-day accuracy and precision range from 4.69% to 13.25% and 1.76% to 6.92%, respectively, for ripretinib. They range from -7.08% to 7.82% and 3.57 to 6.26% for N-desmethyl-ripretinib (**Table 3***, **left***). Inter-day accuracy and precision range from 4.36% to 10.53%, respectively, for ripretinib. They range from -9.06% to 3.05% and 3.08% to 7.39% for N-desmethyl-ripretinib (**Table 3***, **right***).

**Table 3.** Repetitive and intermediate fidelity of ripretinib and N-desmethyl-ripretinib.

| Drugs | Nominal concentration (ng/mL) | Intra-day (n = 5) |  |  | Inter-day (n = 15) |  |  |
| --- | --- | --- | --- | --- | --- | --- | --- |
|  |  | Measured conc. mean ± SD | Precision (%) | Accuracy (%) | Measured conc. mean ± SD | Precision (%) | Accuracy (%) |
| Ripretinib | 50 | 53.1 ± 3.68 | 6.92 | 6.21 | 52.18 ± 3.85 | 7.39 | 4.36 |
|  | 150 | 157.04 ± 8.62 | 5.49 | 4.69 | 156.58 ± 7.86 | 5.02 | 4.39 |
|  | 2500 | 2831.24 ± 49.87 | 1.76 | 13.25 | 2763.21 ± 84.97 | 3.08 | 10.53 |
|  | 4000 | 4336.88 ± 214.79 | 4.95 | 8.42 | 4308.9 ± 183.59 | 4.26 | 7.72 |
| N-desmethyl-ripretinib | 50 | 46.46 ± 1.66 | 3.57 | -7.08 | 45.47 ± 4.11 | 9.04 | -9.06 |
|  | 150 | 142.39 ± 8.92 | 6.26 | -5.07 | 145.23 ± 8.2 | 5.65 | -3.18 |
|  | 2500 | 2695.41 ± 116.41 | 4.32 | 7.82 | 2576.37 ± 125.25 | 4.86 | 3.05 |
|  | 4000 | 4144.3 ± 237.91 | 5.74 | 3.61 | 4093.08 ± 220.5 | 5.39 | 2.33 |

Consequently, all these results ensure that the method has adequate accuracy and precision according to the guidelines criteria of acceptance (17).

#### 2.3.5 Carry-over and dilution integrity

We did not observe a significant carry-over effect in chromatograms acquired upon injection blank sample after injection of the ULOQ (**Table 4**). This result ensures that a patient with a high concentration of ripretinib and/or N-desmethyl-ripretinib does not impact the measurement of the next patient’s sample.

**Table 4.** Carry-over analysis of ripretinib and N-desmethyl-ripretinib.

| Drugs | ULOQ Area ± SD | Blank Area ± SD | LLOQ Area ± SD | %Carry-over ± SD |
| --- | --- | --- | --- | --- |
| Ripretinib | 16756.80 ± 372.22 | 13.40 ± 6.73 | 159.60 ± 21.28 | 8.65 ± 4.77 |
| [<sup>13</sup>C<sub>6</sub>]-ripretinib | 4065.00 ± 84.63 | 4.60 ± 2.97 | 3383.20 ± 260.35 | 0.14 ± 0.09 |
| N-desmethyl-ripretinib | 28206.80 ± 1702.27 | 24.80 ± 8.04 | 283.00 ± 40.35 | 8.81 ± 2.89 |
| [<sup>13</sup>C<sub>6</sub>]-N-desmethyl-ripretinib | 16768.80 ± 906.50 | 19.40 ± 8.08 | 14397.20 ± 1604.89 | 0.14 ± 0.07 |

The accuracy of the diluted samples ranged between 3.90% and 11.12% for ripretinib and -3.35% to 5.50% for N-desmethyl-ripretinib. Furthermore, the overall precision was below 4.5%. These results confirmed the integrity of the dilution process for ripretinib and N-desmethyl-ripretinib, enabling the dilution of patient plasma samples if their concentration exceeded the calibration range (**Table 5**).

**Table 5.** Dilution integrity analysis of ripretinib and N-desmethyl-ripretinib.

| Dilution factor | Nominal conc. (ng/mL) | Ripretinib |  |  | N-desmethyl-ripretinib |  |  |
| --- | --- | --- | --- | --- | --- | --- | --- |
|  |  | Mean ± SD | Precision (%) | Accuracy (%) | Mean ± SD | Precision (%) | Accuracy (%) |
| 1 | 7000 | 7235.31 ± 309.69 | 4.28 | 3.36 | 6383.75 ± 270.14 | 4.23 | -8.80 |
| 2 | 3500 | 3889.22 ± 161.07 | 4.14 | 11.12 | 3692.56 ± 93.69 | 2.54 | 5.50 |
| 10 | 700 | 727.27 ± 29.58 | 4.07 | 3.90 | 676.58 ± 27.64 | 4.09 | -3.35 |

#### 2.3.6 Stability

Overall accuracy and precision were below 13%, ripretinib and N-desmethyl-ripretinib were therefore stable under all storage conditions (**Table 6**). Typically, samples can be extracted after 24 hours if left at room temperature (24°C) or stored at 4°C for up to a week before analysis. In addition, extracted samples remained stable after the post-preparative extraction step under autosampling conditions (15°C).

**Table 6.**
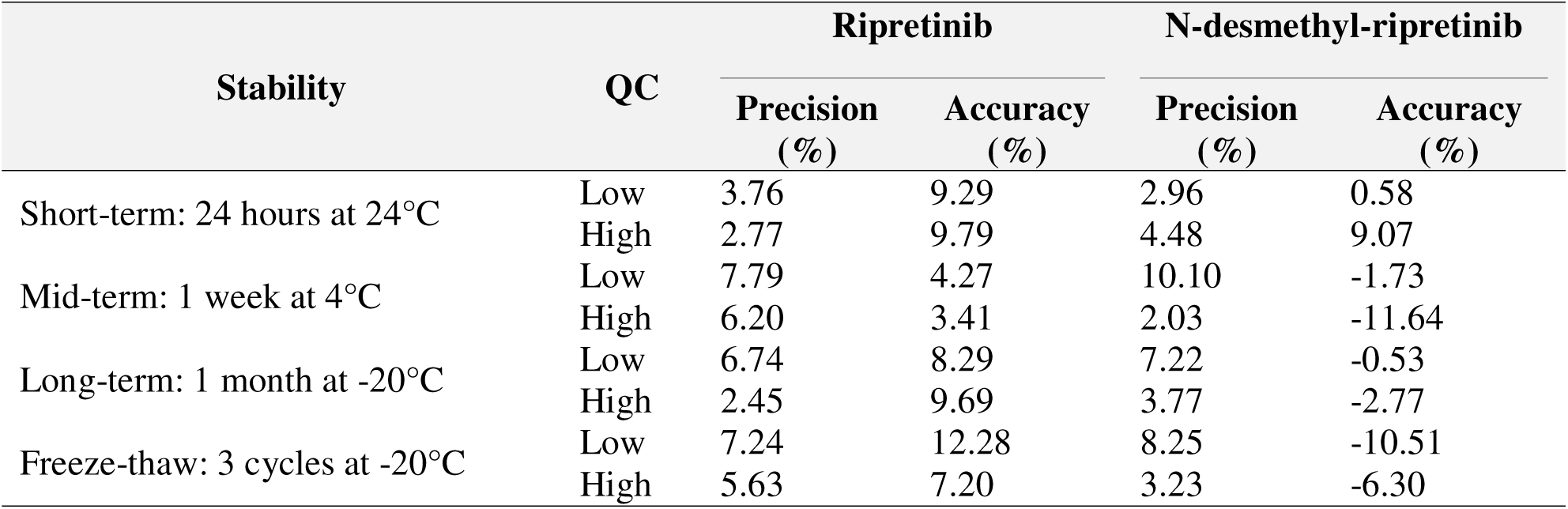
Storage Stability of ripretinib and N-desmethyl-ripretinib.

| Stability | QC | Ripretinib |  | N-desmethyl-ripretinib |  |
| --- | --- | --- | --- | --- | --- |
|  |  | Precision (%) | Accuracy (%) | Precision (%) | Accuracy (%) |
| Short-term: 24 hours at 24°C | Low | 3.76 | 9.29 | 2.96 | 0.58 |
|  | High | 2.77 | 9.79 | 4.48 | 9.07 |
| Mid-term: 1 week at 4°C | Low | 7.79 | 4.27 | 10.10 | -1.73 |
|  | High | 6.20 | 3.41 | 2.03 | -11.64 |
| Long-term: 1 month at -20°C | Low | 6.74 | 8.29 | 7.22 | -0.53 |
|  | High | 2.45 | 9.69 | 3.77 | -2.77 |
| Freeze-thaw: 3 cycles at -20°C | Low | 7.24 | 12.28 | 8.25 | -10.51 |
|  | High | 5.63 | 7.20 | 3.23 | -6.30 |

#### 2.3.7 Reinjection reproducibility

The reproducibility of injection allows us to verify that we obtain consistent and reliable results when injecting at least 5 times a single sample. The accuracy was within the expected range and the precision was < 9% for ripretinib and < 7% for N-desmethyl-ripretinib (**Supplementary Table S2**).

#### 2.3.8 Recovery

We found a recovery in QC samples ranging from 80.93 to 86.93% for ripretinib and from 92.12 to 103.53% for N-desmethyl-ripretinib demonstrating that the extraction method is consistent and reproducible (**Supplementary Table S3**).

### 2.4 Greenness assessment

The sample pretreatment is also external and involves a reduced number of steps. (P1). A volume of 0.05 mL plasma is required (P2) and the measurements are off-line (P3). The procedure involves less than 4 steps (P4) and procedure is manual and involves a miniaturized sample preparation technique (P5). No derivatization agents are involved (P6). The total waste generated is approximately 3.65 g (P7). The number of analytes determined is 2, and the sample throughput is 20 samples per hour (P8). We also used LC-MS/MS (P9). None of the reagents are derived from bio-based sources (P10).

Procedure requires approximately 1.05 mL toxic solvents (P11). Highly flammable and corrosive substances are not avoided (P12). The score for this procedure obtained with AGREE assessment is 0.58 (**Figure 5**), indicating a more sustainable approach compared to other procedures (**Figure 1**).

**Figure 5.**
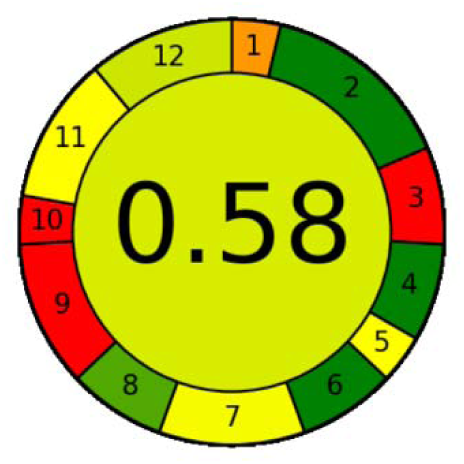
AGREE analysis result for this procedure.

### 2.5 Pharmacokinetic studies

#### 2.5.1 In mice

The plasma concentration-time profile over a 24-hour period, obtained using this LC-MS/MS method, is shown in **Figure 6A**. Mean concentrations (±CV) one hour after oral administration of ripretinib were 567.5 ng/mL (±120.5%) for ripretinib and 183.7 ng/mL (±133%) for its metabolite. The concentrations of the two analytes progressively decreased over time. The mean area under the curve from 0 to 8 hours (AUC_0-8h_) for ripretinib was 2308 ng/mL·h (±86.1%) for oral administration. For N-desmethyl-ripretinib, the mean AUC_0-8h_ value was 973 ng/mL·h (±93.3%). Mice exhibit faster pharmacokinetics and greater inter-individual variability compared to other preclinical models, such as rats (16) or beagle dogs (15) (**Table 7**).

**Figure 6:**
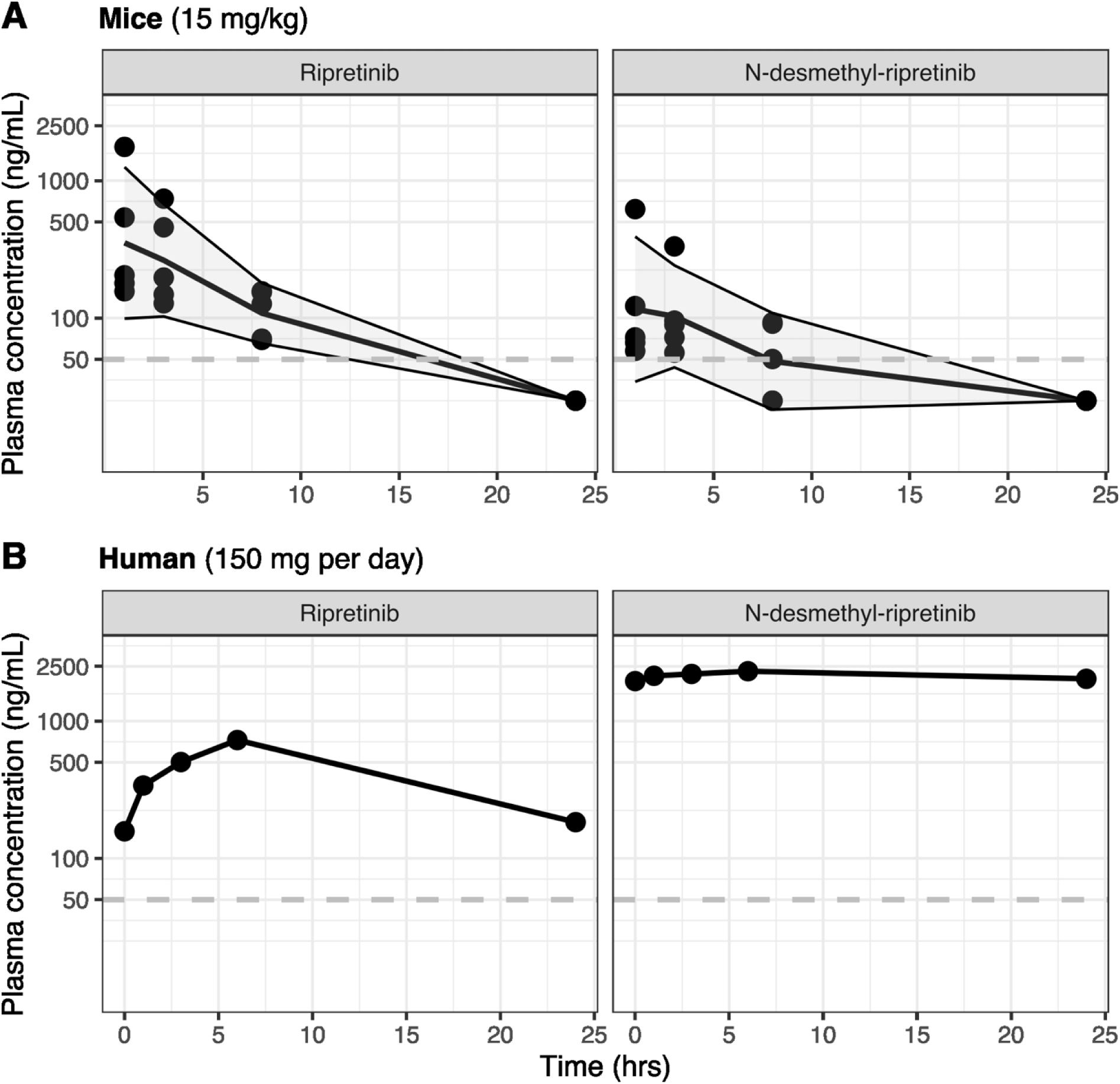
Time-dependent concentration profiles of ripretinib and N-desmethyl-ripretinib. **(A)** Mice were administered a single dose of ripretinib via *per os* (15 mg/kg, n=5). Blood samples were collected at various time points (1, 3, 8 and 24 hours) after administration. All concentration at 24 hours were <LLOQ but are non 0 ng/mL. **(B)** Patient were administered orally once a day dose 150 mg and blood samples were collected just before administration (T0) and at various time points (0, 1, 3, 6 and 24 hours) after administration.

**Table 7.**
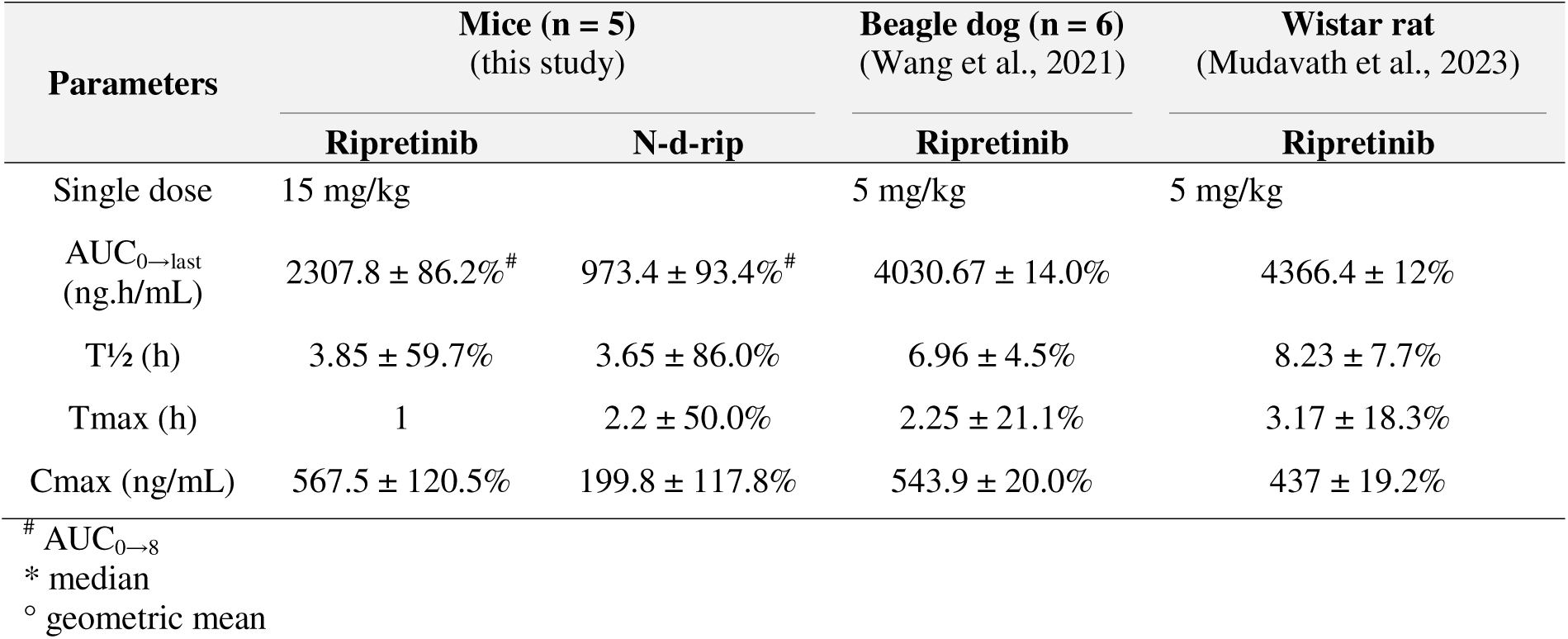
Mean animal pharmacokinetic parameters of ripretinib and N-desmethyl-ripretinib.

#### 2.5.2 On a GIST patient

We determined the pharmacokinetic profile of ripretinib and its metabolite from a GIST patient who was receiving ripretinib at a daily dose of 150 mg in a routine clinical setting (**Figure 6B** and **Table 8**). This patient had several comorbidities that lead to multiple concomitant medications. He did not experience any adverse events The steady-state pharmacokinetic parameters of ripretinib for this patient were: C_max_ = 727.6 ng/mL, T_1/2_ = 9 h, T_max_= 6 h, and AUC_0-24h_ = 11133.6 ng/mL·h. The estimated CL_ss_/F was 13.5 L/h and V/F was 176.7 L. The pharmacokinetic parameters of the metabolite were C_max_ = 2307.8 ng/mL, T_max_ = 6 h, and AUC_0-24h_ = 52203.3 ng/mL·h. The observed pharmacokinetic profile indicated a low elimination of the metabolite within the time interval measured between two drug administrations.

**Table 8.** Mean human pharmacokinetic parameters of ripretinib and N-desmethyl-ripretinib.

| Parameters | Patient (this study) |  | Janku et al., 2020 (n=11) |  | Li et al., 2022 (n = 13) |  | Lin et al., 2025 (n = 4) |  |
| --- | --- | --- | --- | --- | --- | --- | --- | --- |
|  | Ripretinib | N-d-rip | Ripretinib | N-d-rip | Ripretinib | N-d-rip | Ripretinib | N-d-rip |
| Once daily dose | 150 mg |  | 150 mg |  | 150 mg |  | 150 mg |  |
| AUC<sub>0-12</sub> (ng.h/mL) | 6748 | 26716 | 5678°<br>± 32.1% | 7138°<br>± 44.4% |  |  | 9655.2<br>± 21.7% | 9110.7 ± 37.1% |
| T<sub>1/2</sub> (h) | 9 |  |  |  |  |  | 12.5 ± 26.7% | 33.8 ± 65.6% |
| T<sub>max</sub> (h) | 6 | 6 | 2.1* (1-8.1) | 6* | 6* | 6* | 3.6 ± 51.1% | 3.2 ± 73.1% |
| C<sub>max</sub> (ng/mL) | 727.6 | 2307.8 | 761°<br>± 31.8% | 804°<br>± 45.5% | 833*<br>(308-1700) | 1250*<br>(276-1930) | 1088.6<br>± 18.7% | 863.7 ± 38.8% |
| C<sub>min</sub> (ng/mL) | 157.4 | 1954.2 | 284<br>± 62.5% | 546<br>± 78.2% |  |  | 284± 62.5% | 546 ± 78.2% |

\* median
° geometric mean

This was accompanied by high concentrations, likely reflecting an enhanced individual metabolic capacity as evidenced by the measured N-desmethyl-ripretinib /ripretinib AUC_0-24h_ ratio (value of 4.6).

Data available on its pharmacokinetics are scarce (**Table 8**). To our knowledge, available pharmacokinetic data on ripretinib in human are limited to those described in clinical trials (11,24) and in a pharmacokinetic study conducted in four Chinese patients (19). The pharmacokinetics of ripretinib and its equally active metabolite were evaluated following single doses in healthy subjects and multiple doses in patients with advanced malignancies. According to clinical studies, ripretinib and N-desmethyl-ripretinib pharmacokinetics are characterized by a wide inter-patient variability (CV% above 40% for steady-state apparent volume of distribution and apparent clearance). The reported values of plasma maximal concentrations are characterized by a high CV% of 32% and 44%, respectively for ripretinib and its metabolite.

In the light of this pharmacokinetic variability and the possibility of drug dosage increase without toxicity, one hypothesis is TDM would help to optimize treatment response in patients with tumor progression. Moreover, the relationship between ripretinib/ N-desmethyl-ripretinib plasma exposure and treatment efficacy has not been established. Thus, we believe that one major concern would be to explore the variability of pharmacokinetics of ripretinib and its metabolite, in order to further assess the relationship between plasma exposure and clinical response in term of efficacy and/or toxicity.

The quantification of both ripretinib and its metabolite would offer remarkable insights into drug behavior, particularly in polymedicated patients with comorbidities. This is especially relevant for complex clinical cases like our patient.

Indeed, we observed that this patient had exceptionally high concentrations of the metabolite, with plasma concentrations about five times higher than ripretinib itself (metabolic ratio of 4.6). Notably, the metabolite exposure was significantly higher than previously reported in the literature: in patients treated for at least 15 days, Janku et al reported a geometric mean C_min_ (CV%) of 546 ng/mL (± 78.2%) for the metabolite, with a metabolic ratio of 1.29 (± 27.9) (11). In a Chinese patient cohort, the median C_min_ (range) of N-desmethyl-ripretinib was 654.74 ng/mL (range of 30.71-1522.48). In contrast, our patient’s levels were significantly above these reported ranges, despite his C_min_ and C_max_ concentrations of ripretinib falling within expected values. It is well established that coadministration of ripretinib with strong CYP3A4/5 inhibitors are able to markedly increase the concentrations of both ripretinib and its metabolite (25). However, our patient was not receiving any known strong CYP3A4/5 inhibitors, suggesting an intrinsically enhanced metabolic capacity. Moreover, his plasma protein levels exhibited considerable fluctuations over time. Since both ripretinib and its metabolite are highly plasma protein, including serum albumin and α-1 acid glycoprotein, these intra-individual variations of plasma protein levels are likely to influence the free drug levels. In particular, our patient had fluctuating and high levels of α-1 acid glycoprotein consistent with his inflammatory state. This may have increased the free fraction of ripretinib, potentially enhancing its tissue distribution and hepatic metabolism.

Overall, this clinical case and the unexpected pharmacokinetic results observed in this patient highlight the need of further exploring inter-individual variability in ripretinib pharmacokinetics and identifying key covariates that influence drug metabolism and response across various patient populations.

## 3 Conclusion

In conclusion, we developed and validated a rapid, simple and sustainable LC-MS/MS analytical method for the simultaneous quantification of ripretinib and N-desmethyl-ripretinib in plasma. This method is expected to have broad applications in oncology, enabling preclinical studies to better understand the pharmacological effect of ripretinib. Additionally, in GIST patients, it will facilitate further exploration of the relationship between plasma exposure to ripretinib and its active metabolite and clinical response, while also helping to confirm the benefits of therapeutic drug monitoring.

## 4 Materials and Methods

### 4.1 Data and Search Strategy

We source data from the PubMed (https://pubmed.ncbi.nlm.nih.gov) database. We searched for “ripretinib” and “liquid chromatography” to identify publications relating to this method quantification, and “ripretinib” and “pharmacokinetic” for pharmacokinetic data. We selected original research articles published in English up to March 15th, 2025.

### 4.2 Chemical and Reagents

Ripretinib (SVI-ALS-25-020), [^13^C_6_]-ripretinib (PS-ALS-23-005-P4), N-desmethyl-ripretinib (NW- ALS-23-154-P9) and [^13^C_6_]-N-desmethyl-ripretinib (SA-ALS-23-164-P1) were purchased from Alsachim (Illkirch-Graffenstaden, France). Liquid chromatography grade solvents were provided by Prochilab (Paris, France). Human plasmas were obtained from EFS (Etablissement Français du Sang, Bordeaux, France).

### 4.3 Analytical Procedures

#### 4.3.1 Liquid Chromatography and Mass Spectrometry Conditions

Analysis was conducted using an Acquity UltraPerformance Liquid Chromatography (LC) system (Waters, Milford, USA) connected to the MassLynx software. The column was an Acquity Cortecs C18+ 1.6 µm 2.1 × 50 mm column (Waters, Milford, USA). The mobile phase A was distilled water with acetic acid 1% and formic acid 0.1%; the mobile phase B was acetonitrile. The gradient used was as follows: starting with 80% A/ 20% B mobile phases. B was gradually increased to 40% at 0.2 min, 60% at 0.4 min, 80% at 0.6 min, and 90% at 0.8 min. B was maintained at 90% for 0.75 min before being reduced to 20% from 2.1 min until the end of the analysis. The flow rate was 0.3 mL/min. The column temperature was fixed at 25°C, and 7 µL was injected in partial loop mode.

An Acquity TQD detector (Waters, Milford, USA) was used with an electrospray ionization (ESI) source set to positive ion mode, using nitrogen as the nebulization and desolvation gaz. The ESI ion source was operated at 150°C, with the desolvation temperature set to 400°C. The cone gas flow was adjusted to 50 L/h, desolvation flow to 1000 L/h, and the capillary voltage was set at 3.0 kV. The collision in the MS was carried out with argon at a pressure of 3 × 10^-3^ mBar.

#### 4.3.2 Preparation of Standard Stock Solutions, Calibration Standards, and Quality-Control Samples

Individual stock solutions were prepared from powder dissolved in acetonitrile to a final concentration of 1 mg/mL. The calibration solutions of ripretinib and N-desmethyl-ripretinib were mixed and serially diluted in human plasma in the concentration range of 50 – 5000 ng/mL. The calibration standards for the analytical curve were set at concentrations of 50, 100, 250, 750, 2000, 3000, and 5000 ng/mL. Quality control (QC) samples at low (150 ng/mL), medium (2500 ng/mL) and high (4000 ng/mL) concentrations were prepared in plasma. An IS daughter solution was prepared from the stock solution by mixing [^13^C_6_]-ripretinib and [^13^C_6_]-N-desmethyl-ripretinib in acetonitrile mixture and stored at -20°C.

#### 4.3.3 Sample preparation

Blood samples collected in heparinized tubes were centrifuged at 5,000 rpm for 10 minutes to isolate plasma, which was then stored at -20°C until analysis. Plasma samples were thawed at 24°C, and 50 μL of each sample was extracted in 1.5 mL tube using 150 μL of the IS daughter solution prepared in **section 2.4**. The tube was vortexed for 10 seconds and then centrifuged at 10,000 rpm for 5 minutes. The resulting supernatant was collected and directly injected onto the LC-MS analytical platform.

### 4.4 Method Validation

Analytical method validation was performed on human drug-free plasmas spiked with ripretinib and N-desmethyl-ripretinib. We followed the current version of ICH Guidelines on bioanalytical method validation provided by the EMA (17). The validation process includes the following assessed criteria: *matrix effect, selectivity*, *specificity*, *linearity*, *precision and accuracy*, *carry-over*, *dilution integrity*, *stability*, *reinjection reproducibility* and *recovery*. All validation criteria are resumed in **Supplementary table S1**.

Two key aspects of measurement are frequently emphasized: accuracy and precision. Accuracy refers to the degree to which a measurement matches the expected value, whereas precision refers to the degree of consistency among multiple measurements. The formula for the calculation is as follows:

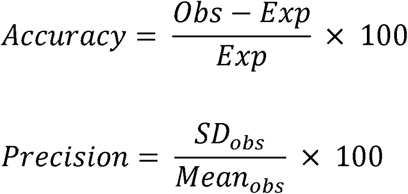

Where *obs* correspond to observed value, *exp* correspond to expected (nominal/theoretical) value and *SD* correspond to standard deviation. In contrast to accuracy, which can be either negative or positive, precision is limited to positive values only.

#### 4.4.1 Matrix effect

The ionization step can be influenced by co-eluted components, which may either enhance or suppress the ionization of the target analyte, leading to changes in the signal detected on the analytical platform. This phenomenon is known as the matrix effect, which requires investigation. To assess the matrix effect, a minimum of six individual samples were prepared, including triplicate sets of low and high concentration quality control samples. Accuracy should be ± 15% and precision should not exceed 15% of the nominal value.

#### 4.4.2 Selectivity and specificity

Selectivity is achieved by distinguishing the analyte from endogenous substances present in blank biological matrices, while specificity is achieved by distinguishing the analyte from exogeneous compounds in complex sample matrices. Selectivity was assessed with at least 6 individual blank samples, and specificity was assessed with at least 6 individual complex samples. A range of drug classes were employed, including PKIs (*i.e.*, bosutinib, dasatinib, encorafenib, imatinib, nilotinib, regorafenib, sunitinib, sorafenib), antiepileptic drugs (*i.e.*, gabapentine, topiramate, lacosamide), corticoids (*i.e.* prednisone, prednisolone), antidepressants (i.e., amitriptyline, citalopram, duloxetine, fluoxetine, hydroxyzine, mianserine, mirtazapine, sertraline), and anti-tuberculosis (ethambutol, isoniazid, pyrazinamide, rifampicine). The abundance attributable of interfering components must be less than 20% of the analyte and less than 5% of the IS observed in the lower limit of quantification (LLOQ).

#### 4.4.3 Linearity

The calibration curve defines the concentration range where the analyte can be accurately measured. It is composed by a blank sample (without any component), a zero sample (only IS) and at least 6 calibrators levels. The lower limit of detection (LOD) was established at three times the signal of background noise, and the lower limit of quantitation (LLOQ) was set at 10 times greater. Linearity refers to the direct relationship between the nominal concentration of the analyte and the signal generated by the analytical platform. The relationship should fit with the simplest regression model. Linearity must be replicated with at least 3 independent runs. Accuracy and precision should be ±20% and less than 20%, respectively, for the LLOQ and ±15% and less than 20%, respectively, at the other levels.

#### 4.4.4 Repetitive and intermediate fidelity

These parameters evaluate intra- and inter-day repeatability. The first parameters were based on the quantification of four levels of quality control (LLOQ, low, middle, and high-level) using quintuplicate extractions in one day, while the second corresponds to the comparison of the first one across three independent days. Accuracy should be ±20% for the LLOQ and 15% for the other levels and precision should not exceed 20% for the LLOQ and 15% for the other levels.

#### 4.4.5 Carry-over and dilution integrity

Carry-over evaluates the persistence of the analyte from a previous sample in the system. Carry-over was performed by analyzing a blank sample directly injected after the highest calibration standard concentration. (5,000 ng/mL for ripretinib and N-desmethyl-ripretinib). The abundance found in the blank sample should not exceed 20% of the analyte abundance in the LLOQ and 5% for the IS. The formula for the calculation of the carry-over is as follows:

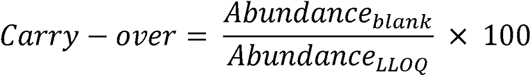

For testing the dilution integrity, a sample spiked at 7,000 ng/mL (exceeding the upper limit of quantification - ULOQ) are diluted in human plasma at 1:2 and 1:10 (v:v). For diluted samples, accuracy should be within ±15% and precision less than 15%.

#### 4.4.6 Stability

The stability analysis corresponds to the ability to maintain consistent and reliable results over time at every stage of preparation and analysis. The stability of compounds was investigated at both low and high levels of QC under various storage conditions: short-term storage (24h at 24°C), mid-term storage (1 week at 4°C), long term storage (1 months at -20°C) and 3 cycles of freeze-thaw at -20°C. Accuracy of ripretinib and N-desmethyl-ripretinib should be ±15% and precision not exceed 15%.

#### 4.4.7 Reinjection reproducibility

A same sample of low, middle and high-level QC were reinjected at least 5 times in the analytical platform. Since the accuracy should be within 15% of the true value, the precision of the measurement should be less than or equal to 15%.

#### 4.4.8 Recovery

Extraction recovery rate is an additional consideration that assesses the efficiency of the extraction process. It was calculated by comparing the abundance obtained from drug-free human plasma samples spiked with analyte of interest before or after the extraction. It was performed on the 3 levels of QCs and should be consistent and reproducible.

### 4.5 Animal and human pharmacokinetic studies

The validated method was applied in a pharmacokinetic study in male RAGγ2C^−/−^ mice. Experimental procedures and protocols were approved by the local ethics committee (agreement number: C335222). Ripretinib was administered orally (15 mg/kg). Heparinized tubes were used to collect 100 µL blood samples in the mandibular vein at different time points (1, 3, 8 h and 24 h) after injection. Samples were centrifuged at 5 000 rpm for 10 minutes, and the supernatant plasma was collected and stored at -20°C until analysis.

Meanwhile, blood samples were collected at different times (0, 1, 3, 6 and 24 h) from a patient who had been receiving ripretinib at the daily dose of 150 mg to treat a GIST in our hospital. Plasma was collected and stored at -20°C until analysis. Institutional review board of Institut Bergonié (Clinical Research College) gave ethical approval for this work. The patient provided informed consent for us to proceed with the sampling and the pharmacokinetic analysis.

### 4.6 Analysis and Graphical Representation

The validation data were analyzed and visualized using R Studio IDE (version 2023.06.0 - http://www.rstudio.com/) with ggplot2 package (26).

The plasma data were fitted in a non-compartmental analysis on Pkanalix software in order to obtain the terminal rate constant (λ_z_) and the area under the concentration- time profiles (AUC_0-last_). The terminal half-live in plasma was calculated from the equation: t_1/2_ = 0.693/λ_z_. The area under the concentration- time profile (AUC_0-last_) was calculated using the trapezoidal method. For the patient, the clearance CL_ss_/F and the volume of distribution Vd/F at the pharmacokinetic steady-state were estimated.

We used the Analytical GREEnness calculator to evaluate the criteria drawn from principles of green analytical chemistry (6).

## Data Availability

All data produced in the present study are available upon reasonable request to the authors

## Acknowledgements

We would like to sincerely thank the medical team of Institut Bergonié: Benoît Rucheton, Maud Toulmonde, Mariella Spalato Ceruso and Florent Peyraud.

## Supplementary Material

See Supplementary material documents

