## Supplementary Figure S1 for "Development and validation of a LC-MS/MS method for ripretinib and its metabolite: example of a journey from laboratory bench to routine application with a greenness assessment"

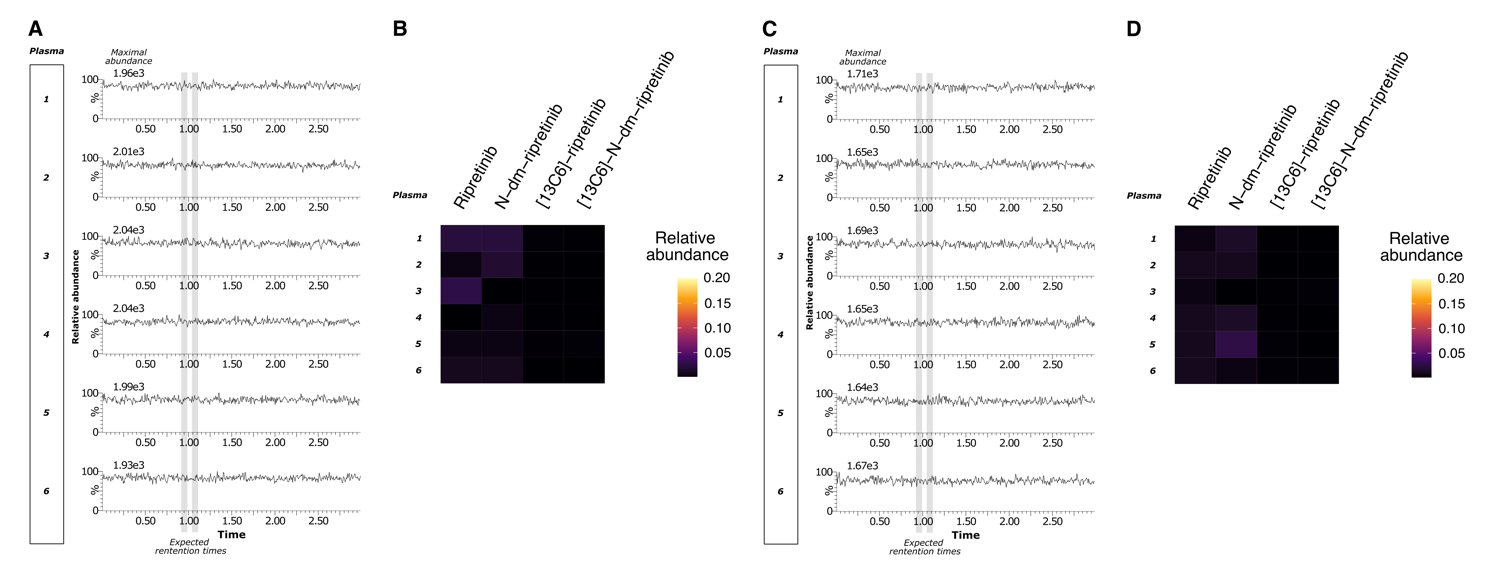

Supplementary Figure S1. Selectivity and Specificity analysis

(**A**) and (**B**) Analysis of individual blank human plasmas. (**A**) Chromatograms of patient plasmas displayed the superposition of MRM channels of ripretinib, N-desmethyl-ripretinib, [13C6]-ripretinib and [13C6]-N-desmethyl-ripretinib. (**B**) Heatmap of the relative abundance of these different blank plasmas relative to the abundance observed in LLOQ samples. (**C**) and (**D**) Analysis of blank plasmas spiked with various compounds (*1 and 2, PKIs ; 3, antiepileptics ; 4, corticoids ; 5, antidepressants ; 6, anti-tuberculosis*). (**D**) Heatmap of the relative abundance of these different spiked plasmas relative to the abundance observed in LLOQ samples. For the interpretation of heatmaps, warm colors represent the abundance of patient plasma concentrations near or exceeding 20% of the LLOQ abundance, while dark colors signify abundance near 0%.
