## Supplementary Figure S2 for "Development and validation of a LC-MS/MS method for ripretinib and its metabolite: example of a journey from laboratory bench to routine application with a greenness assessment"

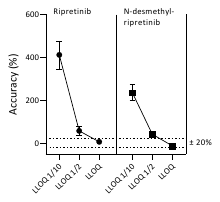

Supplementary Figure S2. LLOQ dilution

Graph illustrating the accuracy of dilutions at half and one-tenth of the LLOQ (n=5).
