## Supplemental Table S1 for "Development and validation of a LC-MS/MS method for ripretinib and its metabolite: example of a journey from laboratory bench to routine application with a greenness assessment"

Supplementary Table S1. EMA validation criteria

| Criteria | Definition | Elements | Acceptance |
| --- | --- | --- | --- |
| Selectivity | Ability to discriminate the analyte of interest from potential interfering substance in the blank biological matrix | ≥ 6 individual blank samples | Abundance attributable of interfering components <20% of the analyte in the LLOQ  Abundance attributable of interfering components <5% of the IS in the LLOQ |
| Specificity | Ability to accurately detect an analyte in the presence of other compounds in a complex sample matrix |  |  |
| Matrix effect | Interference on analyte signal caused by co-eluted matrix components on the analytical platform performance | ≥ 6 individual samples  ≥ 3 replicates of 2 QCs : low and high | Precision should not exceed 15%  Accuracy should be ±15% of the nominal concentration |
| Calibration curve | Relationship between the theoretical (nominal) concentration of an analyte and the signal produced (response) by a measuring instrument | Same biological matrix  re  Relationship should fit with the simplest regression model  ≥ 3 Independent runs | Accuracy should be ±20% at the LLOQ and ±15% at the other levels  ≥ 75% of the calibrators with ≥ 6 calibrators levels should meet the above criteria |
| Accuracy & Precision | Closeness of a measured value to the nominal value & degree of consistency and reproducibility of a set of measurements. | ≥ 5 replicates of 4 QCs : LLOQ, low, middle and high  ≥ 3 Independent runs | Precision should not exceed 20% for LLOQ and 15% for the other levels  Accuracy should be ±20% of the LLOQ value and ±15% for the other levels |
| Carry-over | Persistence of an amount of analyte from a previous sample in the analytical instrument | Injection of blank sample after high concentration sample (*e.g.* ULOQ) | Abundance of blank sample after ULOQ <20% of the analyte in the LLOQ  Abundance of blank sample after ULOQ <5% of the IS in the LLOQ |
| Dilution integrity | Evaluation of sample dilution procedure | ≥ 5 replicates of a QC > ULOQ, pure and per dilution factors of this QC | Precision should not exceed 15%  Accuracy should be ±15% of the nominal concentration |
| Stability | Ability to maintain consistent and reliable results over time at every stage of preparation and analysis | ≥ 3 replicates of 2 QCs : low and high  Different storage conditions:   - Freeze-thaw stability in matrix (≥ 3 cycles at -20°C/-80°C) - Short-term stability (X hours/days at bench-top temperature) - Long-term stability (Y days at -20°C/-80°C) - Processed sample stability (e.g. extract samples, samples in autosampler) | Accuracy should be ±15% of the nominal concentration |
| Reinjection reproducibility | Ability to obtain consistent and reliable results when the same sample is injected multiple times | ≥ 5 replicates of 3 QCs : low, middle and high | Precision should not exceed 15%  Accuracy should be ±15% of the nominal concentration |
| Recovery* | Efficiency of extraction | ≥ 3 replicates of 3 QCs : low, middle and high *vs* blanks spiked with the analyte | Doesn't have to be 100% (analyte + IS), the extent of recovery must be consistent and reproducible |

***Abbreviations: IS****, internal standard;* ***LLOQ****, lower limit of quantification;* ***QC****, quality control;* ***ULOQ****, upper limit of quantification*

**Additional considerations*
