## Supplementary Table S2 for "Development and validation of a LC-MS/MS method for ripretinib and its metabolite: example of a journey from laboratory bench to routine application with a greenness assessment"

Supplementary Table S2. Reinjection reproducibility

| **QC** | **Nominal conc. (ng/mL)** | **Ripretinib** | | | **N-desmethyl-ripretinib** | | |
| --- | --- | --- | --- | --- | --- | --- | --- |
|  |  | **Mean ± SD** | **Precision (%)** | **Accuracy (%)** | **Mean ± SD** | **Precision (%)** | **Accuracy (%)** |
| Low | 150 | 155.92 ± 12.93 | 8.29 | 3.95 | 145.66 ± 10.15 | 6.97 | -2.90 |
| Mid | 2500 | 2609.88 ± 37.13 | 1.42 | 4.40 | 2316.74 ± 32.78 | 1.41 | -7.33 |
| High | 4000 | 4488.11 ± 98.64 | 2.20 | 12.20 | 4045.80 ± 114.06 | 2.82 | 1.15 |
