## Supplementary Table S3 for "Development and validation of a LC-MS/MS method for ripretinib and its metabolite: example of a journey from laboratory bench to routine application with a greenness assessment"

Supplementary Table S3. Extraction recovery analysis

| **QC** | **Ripretinib** | | | **N-desmethyl-ripretinib** | | |
| --- | --- | --- | --- | --- | --- | --- |
|  | **Area in pure solution ± SD** | **Area after extraction ± SD** | **Extraction recovery ± SD** | **Area in pure solution ± SD** | **Area after extraction ± SD** | **Extraction recovery ± SD** |
| Low | 655.5 ± 77.42 | 530.5 ± 38.10 | 80.93 ± 5.81 | 672.0 ± 100.63 | 695.75 ± 59.67 | 103.53 ± 8.88 |
| High | 18768.5 ± 2143.1 | 16315.0 ± 624.7 | 86.93 ± 3.33 | 25581.6 ± 3731.18 | 23566.2 ± 3185.85 | 92.12 ± 10.88 |
